# The association between influenza vaccination and the risk of SARS-CoV-2 infection, severe illness, and death: a systematic review of the literature

**DOI:** 10.1101/2020.09.25.20201350

**Authors:** Marco Del Riccio, Chiara Lorini, Guglielmo Bonaccorsi, John Paget, Saverio Caini

**Affiliations:** Postgraduate Medical School in Public Health, University of Florence, Florence, Italy; Department of Health Sciences, University of Florence, Florence, Italy; Netherlands Institute for Health Services Research (Nivel), Utrecht, the Netherlands; Molecular and Lifestyle Epidemiology Branch, Institute for Cancer Research, Prevention and Clinical Network (ISPRO), Florence, Italy

**Keywords:** Influenza vaccine, SARS-CoV-2, risk, infection, severity, systematic review

## Abstract

We reviewed the association between seasonal influenza vaccination and the risk of SARS-CoV-2 infection or complicated illness or poor outcome (e.g. severe disease, need for hospitalization or ventilatory support, or death) among COVID-19 patients. None of the studies that were reviewed (n=12) found a significant increase in the risk of infection or in the illness severity or lethality, while some reported significantly inverse associations. Our findings support measures aimed at raising influenza vaccination coverage in the coming months.

## Manuscript text

There has been an important debate recently in the media and scientific community and in the media about the relationship between influenza vaccination and COVID-19. Influenza and COVID-19 are respiratory viral illnesses that may be clinically indistinguishable and tend to be life-threatening in largely overlapping population subgroups (e.g. the elderly and people suffering from chronic health conditions). Moreover, being respiratory virus illnesses, their peak of activity may occur in the same period of the year (i.e. winter months in temperate countries). Based on the above considerations, most health professionals have advocated in favour of strengthening influenza vaccination programmes, arguing that rising vaccine coverage could help improve COVID-19 patient management by allowing easier differential diagnosis and reducing the overload of the healthcare systems, particularly intensive care units (ICUs) [1]. Public health decisions should be based as much as possible on the best available evidence regarding any benefits and drawbacks that the proposed intervention may be expected to entail, and since the influenza season is fast approaching in the northern hemisphere, a literature review on this topic is urgently needed. Here, we conducted a systematic review of the articles that examined whether influenza vaccination affects the risk of being infected with the SARS-CoV-2 virus, and the risk of complicated illness or poor outcome (e.g. severe disease, need for hospitalization or ventilatory support, or death) among COVID-19 patients.

The literature search was performed on 31 August, 2020, by interrogating the MEDLINE, Embase and medRxiv databases for both peer-reviewed and non-peer-reviewed articles using the following string: ‘SARS-CoV-2 OR COVID’ AND ‘influenza OR flu AND ‘vaccin*’. All identified articles were first independently screened by two researchers (MDR and SC) based on their title, and any considered as potentially eligible for inclusion by either researcher was then obtained and read in full text. Any disagreement on the eligibility of a given article was resolved by consensus. The literature search was then extended to the reference lists of all the articles that were obtained in full copy (regardless of their final inclusion in the review). To be eligible for inclusion, an article had to be an original report based on individual-level data; studies relying on aggregated data (e.g. ecological studies reporting correlations [2-3]) were not retained because of their higher proneness to bias and confounding. Letters and commentaries with no original data were discarded as well.

The literature search identified 1,619 non-duplicate entries, of which 1,461 were excluded based on their title. The remaining 158 articles were read in full copy, and an additional article was identified in their reference lists. Finally, twelve independent articles met all inclusion criteria and were retained (Figure 1). Seven articles [6-12] focused on the association between influenza vaccination and the risk of SARS-CoV-2 infection (Table 1): these encompassed a total of 242,323 subjects, of which 56.6% were contributed by Pawlowski et al. [11] and 32.6% by Vila-Córcoles A et al. [12]. Most studies were based on subjects from the general population, except the two smallest studies, which included 203 firefighters and paramedics from the USA [7], and 640 liver transplant patients from Italy [8]. The laboratory diagnostic method varied across studies, with (RT-)PCR being used in three of them. No statistical adjustment was made in four studies: Aziz et al. [6] and Donato et al. [8] found no significant association, while COVID-19 cases were significantly less likely to be vaccinated than test-negative subjects in the studies by Jehi et al. [9] (that separately reported on two independent cohorts) and Caban Martinez et al. [7]. Three studies reported measures of association adjusted by age, gender, comorbidities, and other potential confounders, all of which found that influenza vaccinees were significantly less likely to get infected with the SARS-CoV-2 virus than non-vaccinees. Noale et al. [10] found a reduced risk among subjects aged less than 65 years (odds ratio [OR] 0.85, 95% confidence intervals [CI] 0.74-0.98, p=0.024; the OR among those aged ≥65 years was of similar magnitude, 0.87, but not statistically significant, p=0.483). On the contrary, the association was stronger in the ≥65 years subgroup (relative risk 0.74, 95%CI 0.61-0.89, p<0.01) in the study by Pawlowski et al. [11], while achieving statistical significance (p<0.03) in the whole population as well. Finally, a statistically significant inverse association (hazard ratio 0.63, 95%CI 0.43-0.92, p=0.015) emerged among adults aged ≥50 years enrolled in the study by Vila-Córcoles et al. [12].

**Table 1.**
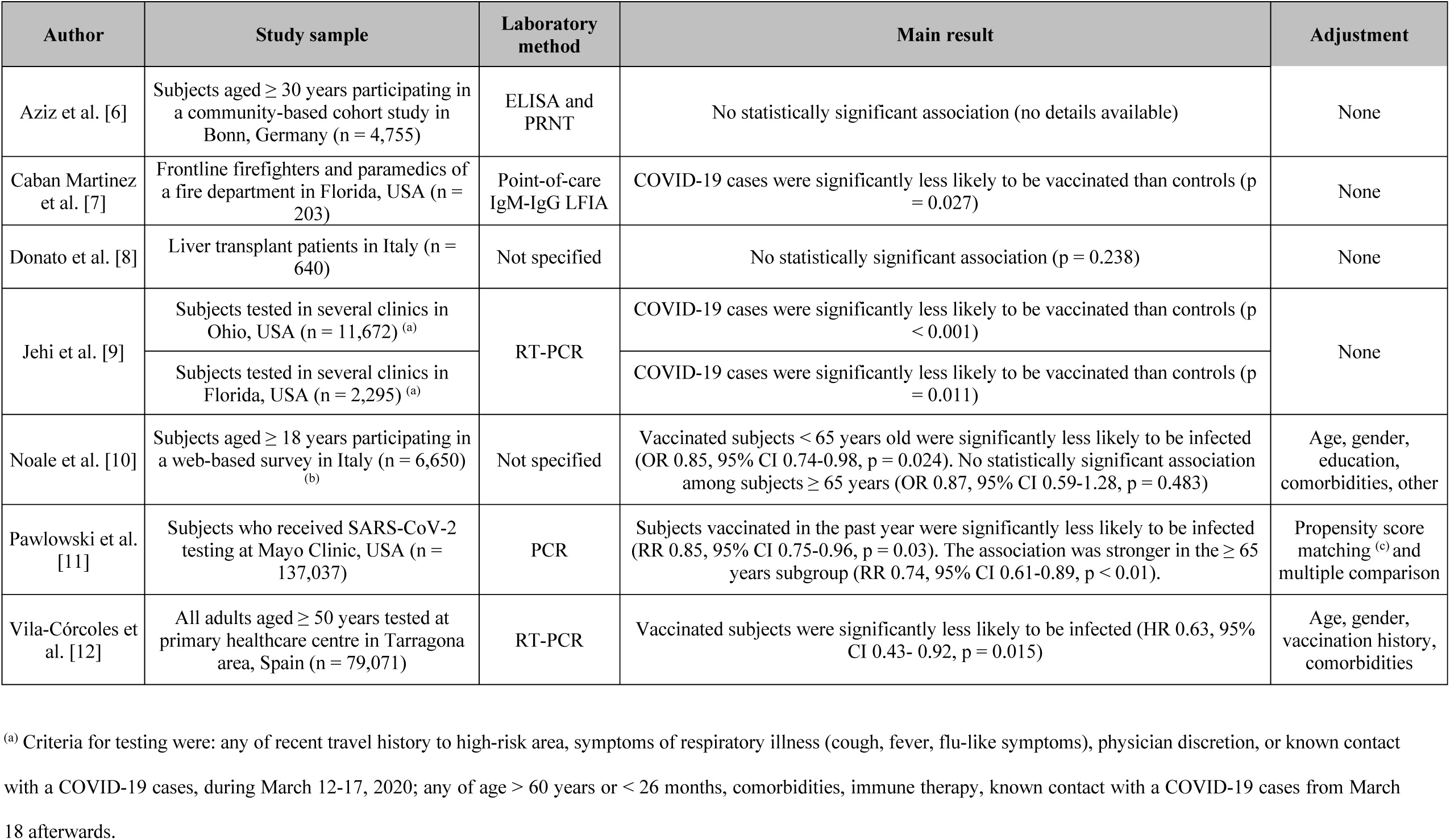
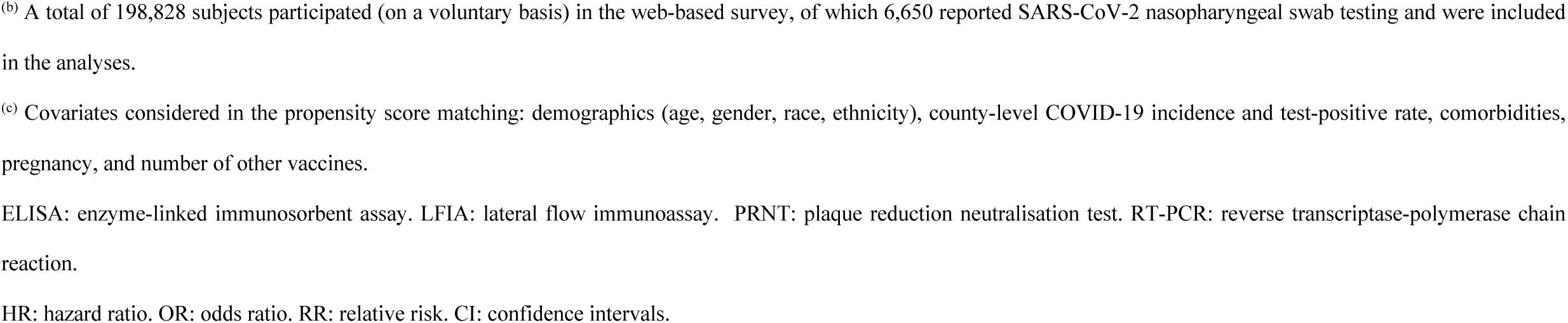
Main features and results of studies on the association between influenza vaccination and the risk of infection with SARS-Cov-2.

| Author | Study sample | Laboratory method | Main result | Adjustment |
| --- | --- | --- | --- | --- |
| Aziz et al. [6] | Subjects aged $\geq 30$ years participating in a community-based cohort study in Bonn, Germany (n = 4,755) | ELISA and PRNT | No statistically significant association (no details available) | None |
| Caban Martinez et al. [7] | Frontline firefighters and paramedics of a fire department in Florida, USA (n = 203) | Point-of-care IgM-IgG LFIA | COVID-19 cases were significantly less likely to be vaccinated than controls (p = 0.027) | None |
| Donato et al. [8] | Liver transplant patients in Italy (n = 640) | Not specified | No statistically significant association (p = 0.238) | None |
| Jehi et al. [9] | Subjects tested in several clinics in Ohio, USA (n = 11,672) <sup>(a)</sup> | RT-PCR | COVID-19 cases were significantly less likely to be vaccinated than controls (p < 0.001) | None |
|  | Subjects tested in several clinics in Florida, USA (n = 2,295) <sup>(a)</sup> |  | COVID-19 cases were significantly less likely to be vaccinated than controls (p = 0.011) |  |
| Noale et al. [10] | Subjects aged $\geq 18$ years participating in a web-based survey in Italy (n = 6,650) <sup>(b)</sup> | Not specified | Vaccinated subjects < 65 years old were significantly less likely to be infected (OR 0.85, 95% CI 0.74-0.98, p = 0.024). No statistically significant association among subjects $\geq 65$ years (OR 0.87, 95% CI 0.59-1.28, p = 0.483) | Age, gender, education, comorbidities, other |
| Pawlowski et al. [11] | Subjects who received SARS-CoV-2 testing at Mayo Clinic, USA (n = 137,037) | PCR | Subjects vaccinated in the past year were significantly less likely to be infected (RR 0.85, 95% CI 0.75-0.96, p = 0.03). The association was stronger in the $\geq 65$ years subgroup (RR 0.74, 95% CI 0.61-0.89, p < 0.01). | Propensity score matching <sup>(c)</sup> and multiple comparison |
| Vila-Córcoles et al. [12] | All adults aged $\geq 50$ years tested at primary healthcare centre in Tarragona area, Spain (n = 79,071) | RT-PCR | Vaccinated subjects were significantly less likely to be infected (HR 0.63, 95% CI 0.43- 0.92, p = 0.015) | Age, gender, vaccination history, comorbidities |

<sup>(a)</sup> Criteria for testing were: any of recent travel history to high-risk area, symptoms of respiratory illness (cough, fever, flu-like symptoms), physician discretion, or known contact with a COVID-19 cases, during March 12-17, 2020; any of age > 60 years or < 26 months, comorbidities, immune therapy, known contact with a COVID-19 cases from March 18 afterwards.
<sup>(b)</sup> A total of 198,828 subjects participated (on a voluntary basis) in the web-based survey, of which 6,650 reported SARS-CoV-2 nasopharyngeal swab testing and were included in the analyses.
<sup>(c)</sup> Covariates considered in the propensity score matching: demographics (age, gender, race, ethnicity), county-level COVID-19 incidence and test-positive rate, comorbidities, pregnancy, and number of other vaccines.
ELISA: enzyme-linked immunosorbent assay. LFIA: lateral flow immunoassay. PRNT: plaque reduction neutralisation test. RT-PCR: reverse transcriptase-polymerase chain reaction.
HR: hazard ratio. OR: odds ratio. RR: relative risk. CI: confidence intervals.

**Figure 1.**
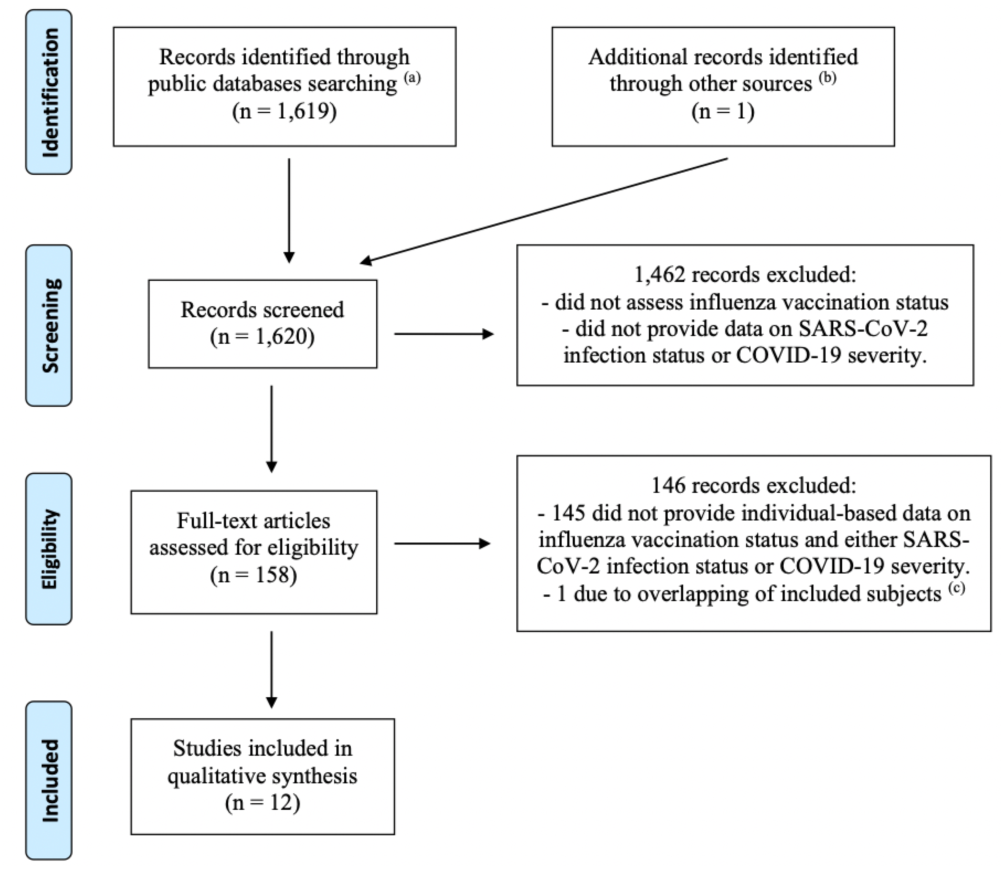
Flow-chart of the literature review (up to August 31, 2020) of studies investigating the association between influenza vaccination status and either the risk of being infected with the SARS-CoV-2 virus, or the risk of severe illness or death among COVID-19 patients ^(a)^ MEDLINE, Embase, MedRxiv ^(b)^ Reference list of records found through public databases ^(c)^ The study by Santos et al. [4] was excluded due to probable overlap of the study population with the article by Fink et al. [5]. Both studies were based on data from hospitalizations for COVID-19 patients in Brazil registered in a national surveillance system. The study by Santos et al. was less recent and based on a smaller population, and examined fewer outcomes, compared to Fink et al, and, unlike the latter, performed no statistical adjustments.

Five articles [5,13-16] reported on the association between influenza vaccination and the risk of severe illness and/or death among COVID-19 patients (Table 2). The total number of patients was 111,820 of which the majority (82.9%) were contributed by Fink et al. [5]. The latter was the only study encompassing a minority (16%) of non-laboratory-confirmed patients. Jehi et al. [13] and Murillo-Zamora et al. [14] found that the likelihood of being vaccinated against influenza was significantly (p<0.001) or, respectively, nearly significantly (p=0.073) lower among patients who required to be hospitalized compared to those who did not. Likewise, Fink et al. [5] reported a significantly lower odds of requiring intensive care or respiratory support among influenza vaccinees vs. non-vaccinees. The latter study also found that vaccinated COVID-19 patients were at significantly reduced risk of dying compared to non-vaccinated ones (OR 0.82, 95%CI 0.75-0.89, p<0.01), but this finding was not confirmed in the studies by Ortiz-Prado et al. [15] and Poblador-Plou et al. [16].

**Table 2.**
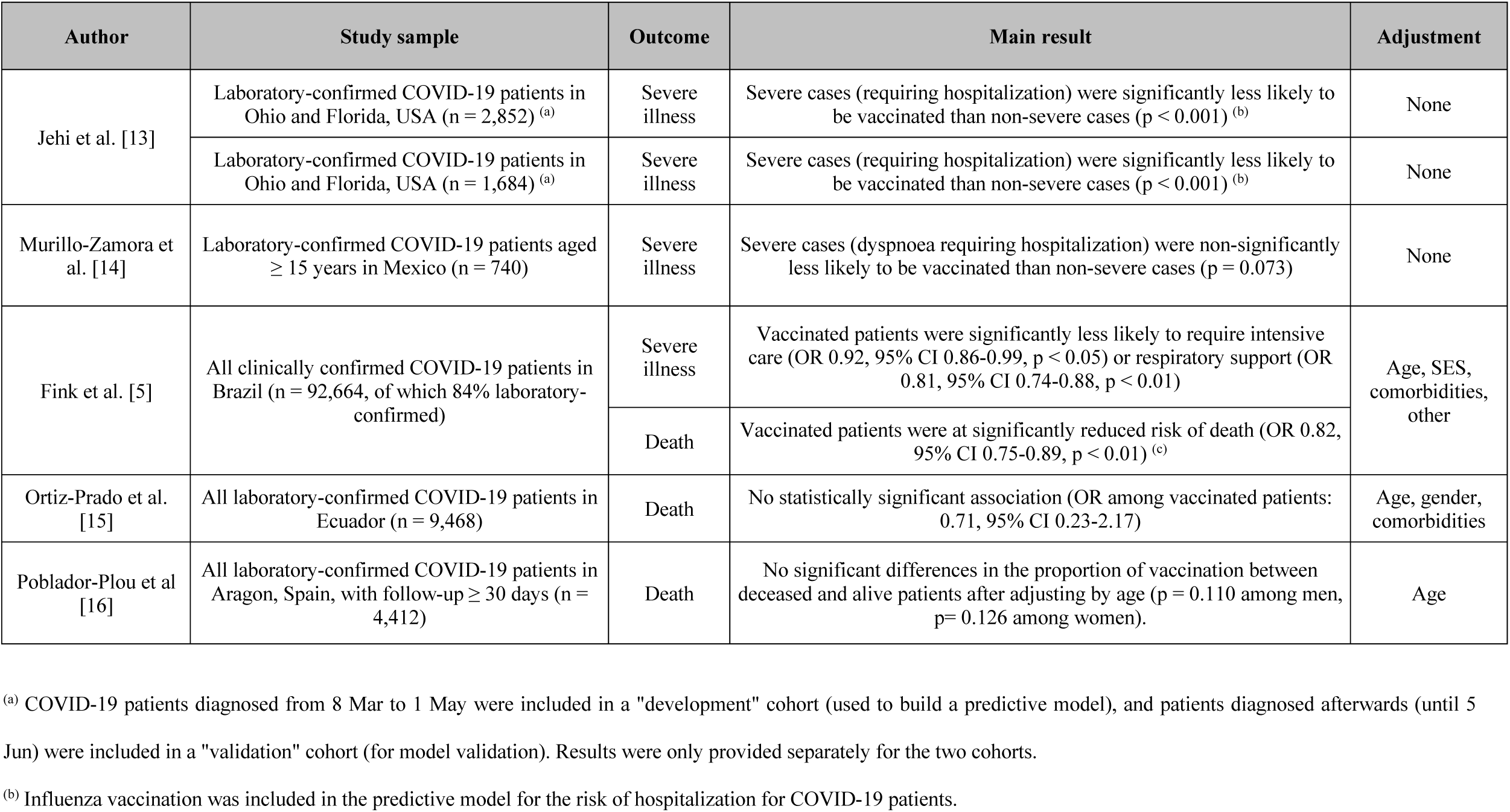
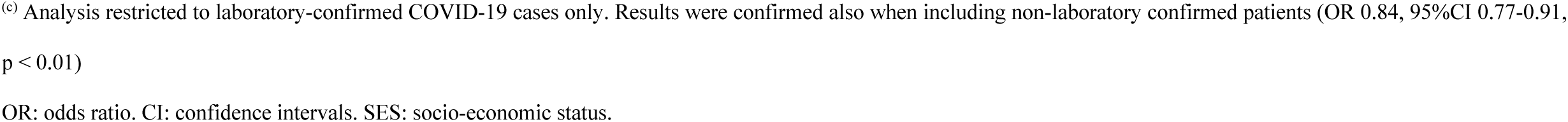
Main features and results of studies on the association between influenza vaccination and the risk of severe illness or death among COVID-19 patients.

| Author | Study sample | Outcome | Main result | Adjustment |
| --- | --- | --- | --- | --- |
| Jehi et al. [13] | Laboratory-confirmed COVID-19 patients in Ohio and Florida, USA (n = 2,852) <sup>(a)</sup> | Severe illness | Severe cases (requiring hospitalization) were significantly less likely to be vaccinated than non-severe cases (p < 0.001) <sup>(b)</sup> | None |
|  | Laboratory-confirmed COVID-19 patients in Ohio and Florida, USA (n = 1,684) <sup>(a)</sup> | Severe illness | Severe cases (requiring hospitalization) were significantly less likely to be vaccinated than non-severe cases (p < 0.001) <sup>(b)</sup> | None |
| Murillo-Zamora et al. [14] | Laboratory-confirmed COVID-19 patients aged ≥ 15 years in Mexico (n = 740) | Severe illness | Severe cases (dyspnoea requiring hospitalization) were non-significantly less likely to be vaccinated than non-severe cases (p = 0.073) | None |
| Fink et al. [5] | All clinically confirmed COVID-19 patients in Brazil (n = 92,664, of which 84% laboratory-confirmed) | Severe illness | Vaccinated patients were significantly less likely to require intensive care (OR 0.92, 95% CI 0.86-0.99, p < 0.05) or respiratory support (OR 0.81, 95% CI 0.74-0.88, p < 0.01) | Age, SES, comorbidities, other |
|  |  | Death | Vaccinated patients were at significantly reduced risk of death (OR 0.82, 95% CI 0.75-0.89, p < 0.01) <sup>(c)</sup> |  |
| Ortiz-Prado et al. [15] | All laboratory-confirmed COVID-19 patients in Ecuador (n = 9,468) | Death | No statistically significant association (OR among vaccinated patients: 0.71, 95% CI 0.23-2.17) | Age, gender, comorbidities |
| Poblador-Plou et al [16] | All laboratory-confirmed COVID-19 patients in Aragon, Spain, with follow-up ≥ 30 days (n = 4,412) | Death | No significant differences in the proportion of vaccination between deceased and alive patients after adjusting by age (p = 0.110 among men, p = 0.126 among women). | Age |

<sup>(a)</sup> COVID-19 patients diagnosed from 8 Mar to 1 May were included in a "development" cohort (used to build a predictive model), and patients diagnosed afterwards (until 5 Jun) were included in a "validation" cohort (for model validation). Results were only provided separately for the two cohorts.
<sup>(b)</sup> Influenza vaccination was included in the predictive model for the risk of hospitalization for COVID-19 patients.
<sup>(c)</sup> Analysis restricted to laboratory-confirmed COVID-19 cases only. Results were confirmed also when including non-laboratory confirmed patients (OR 0.84, 95%CI 0.77-0.91, $p < 0.01$ )
OR: odds ratio. CI: confidence intervals. SES: socio-economic status.

Influenza epidemics recur every year and the persistence of SARS-CoV-2 circulation in the upcoming months may expose healthcare systems to a severe risk of resources scarcity. Influenza vaccination is the cornerstone of influenza prevention, thus attaining higher vaccine coverage has been widely acknowledged as a public health priority [17]. Concerns about a possible association between influenza vaccination and the risk of coronavirus infection were raised based on the Wolff’s paper, which examined endemic coronaviruses circulating in the USA in the 2017-18 season, much earlier the emergence of the SARS-CoV-2 virus [18]. After reviewing the existing literature on the topic, we can safely conclude that influenza vaccination is unlikely to be associated with an increase in SARS-CoV-2 risk of infection or with COVID-19 severity and the risk of associated death. In fact, most reviewed studies detected an inverse relation, which was unexpected and even disconcerting given that influenza vaccines are not designed to protect from SARS-CoV-2. It must be acknowledged, however, that all reviewed studies are retrospective and observational in nature, thus likely to be not devoid of bias, which should warn against drawing premature conclusions regarding this finding. In conclusion, our review finds that based on our knowledge (up to the end of August 2020), public health measures aimed at raising the influenza vaccine coverage should be encouraged and there is no evidence to suggest that this would have a negative impact on populations in terms of SARS-CoV-2 related infections, illness or deaths.

## Data Availability

The data that support the findings of this study are available in the MEDLINE, Embase and medRxiv databases.

